# Estimating transmission pathways of COVID-19 in U.S. nursing homes

**DOI:** 10.1101/2025.09.16.25335445

**Authors:** Jessica M Healy, Prabasaj Paul, Brajendra Singh, Nimalie Stone, Kara Jacobs-Slifka, Rachel B Slayton

## Abstract

**Background:** Pathogen transmission dynamics in nursing homes are unique due to the interactions between staff and residents. Understanding the differences in transmission rates between and among staff and residents can identify the transmission pathways that contributed most to the spread of SARS-CoV-2 in U.S. nursing homes during the pandemic.

**Methods:** We used publicly available U.S. National Healthcare Safety Network (NHSN) surveillance data on resident and staff cases, vaccination, and resident deaths during June 2020–June 2022, to estimate SARS-CoV-2 transmission between and among residents and staff. We used a Bayesian inversion transmission model to produce the estimates. We categorized nursing homes by the number of beds, average length of stay, or community social vulnerability. We estimated weekly transmission events for three periods: pre-Delta (June 2020—March 2021), Delta (April 2021—October 2021), and Omicron (November 2021—June 2022).

**Results:** Weekly transmission events within each nursing homes category were highest between residents in homes with >299 beds during Omicron (0.66, 95% Credible Interval (CrI): 0.13-0.93), from staff to residents in homes with average length of stay >10 weeks during pre-Delta (0.88, 95% CrI: 0.06-1.85), and between residents in homes in communities with the lowest social vulnerability index during pre-Delta (0.89, 95% CrI: 0.07-1.16).

**Conclusions:** Transmission from staff to residents and between residents may have contributed more to SARS-CoV-2 spread in nursing homes than resident-to-staff and staff-to-staff transmission. Interventions interrupting these transmission pathways and that consider community exposures could reduce the size or duration of future outbreaks.

## INTRODUCTION

Pathogen transmission dynamics are unique within nursing homes (NH) due in part to the resident population’s characteristics and their interactions with healthcare staff. Understanding the transmission pathways (e.g., from resident to resident, staff to resident, etc.) that take place in this setting, as well as how transmission varies across resident characteristics and interactions, allows identification of pathways for intervention. During the COVID-19 pandemic, the Centers for Medicare & Medicaid Services required NH to report cases, deaths, and immunizations among residents and staff to the Centers for Disease Control and Prevention’s National Healthcare Safety Network (NHSN) (1). With this information, estimating the amount and directionality of NH transmission for SARS-CoV-2 between and among staff and residents is possible.

Several NH-specific and sociodemographic risk factors for COVID-19 occurrence and severe outcomes in NH and residents have been identified since the pandemic began. These factors included but were not limited to NH with more than 150 beds, location in an urban area, and having a higher proportion of Black and Hispanic residents (2, 3). Given that this analysis was done with national data, we chose to examine the variability in transmission by categorizing NH by size, average length of stay (LOS) among residents, and sociodemographic characteristics of the surrounding county’s population. We estimated SARS-CoV-2 transmission between and among staff and residents by modeling four separate pathways for transmission between and among the two groups using surveillance data in a Bayesian inversion compartmental model for each of the categorizations and outbreak time periods determined by the dominant circulating variant.

## METHODS

### Data sources

Weekly COVID-19 cases, vaccinations, and deaths among residents, weekly cases and vaccinations among NH staff, and the total number of staff working and beds filled each week during June 2020—June 2022 were used from the NHSN for fitting the model (1). NH staff, as defined by NHSN, included employees, consultants, contractors, and volunteers who provided care and services to residents. The total number of staff working per week was missing for the first year in the data (NH staff totals were only required to be reported beginning in June 2021) and in 4% of the subsequent reported facility-weeks. We imputed missing staff counts by multiple imputation chained equations (MICE) using NH identifier, time, location, and NH size as predictors(4). Community COVID-19 data on weekly case prevalence and weekly vaccination rates among those ≥65 years in the county where NH were located were used as parameters for the proportion of admissions that were infected, the proportion vaccinated but not immune (within 4 weeks of vaccination date), or vaccinated and immune. For the purposes of this study, “vaccinated” refers to completion of the two-dose primary vaccine series and “immune” denotes when an individual is at least 4 weeks from the completion of their primary vaccine series or infection. All NH-level data were averaged to weekly counts to represent an average NH, and separate models were constructed for each period of SARS-CoV-2 variant dominance (i.e., pre-Delta: June 1, 2020—March, 31 2021, Delta: April 1, 2021—October 31, 2021, Omicron: November 1, 2021—June 5, 2022).

#### Nursing home categorization

Multiple categorization schemes were used to aggregate the data to produce transmission rate estimates for representative NH types. NH size, average resident LOS, and population characteristics for the county where NH were located were categorized, and rate estimates were produced for each. For NH size, homes were divided between those with at least 300 beds, representing the largest 1% of NH, and those with less than 300 beds, given their difference in incidence patterns. For NH-wide LOS, the tertiles of the median resident LOS were used to define the categories: <6 weeks, 6-10 weeks, and >10 weeks. The CDC Social Vulnerability Index (SVI) was used for county-level population indicators and divided into quartiles (5). The themes and social factors considered in the SVI include the county’s socioeconomics, demographics (including race and ethnicity), housing, and transportation.

### Compartmental model structure

Our model included two groups of compartments, residents and staff, which were connected through the number of cases contributing to transmission in the other group. For the resident group, a SEIVRD (<u>S</u>usceptible, <u>E</u>xposed but not infectious, Infectious, <u>V</u>accinated but not immune, immune through infection or vaccination (<u>R</u>), <u>D</u>eath) model framework was constructed (Fig. 1), and for the staff group, a SEIVR model framework was used (Fig. 2). All compartments were scaled by the weekly number of residents or staff reported by the NH, producing proportions. All vacancies in NH due to deaths and discharges among residents were replaced with new admissions from the community (Fig. 2).

**Figure 1.**
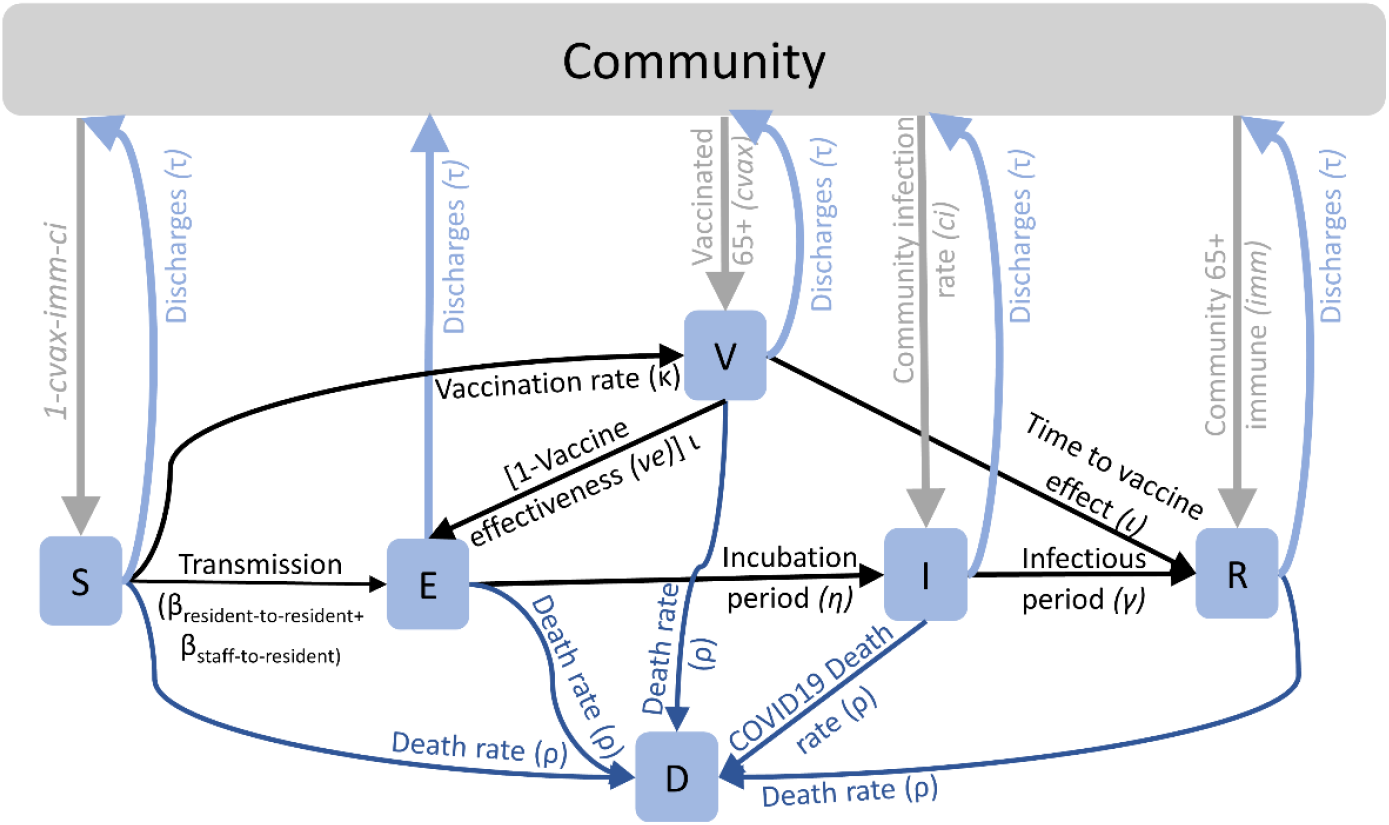
Schematic of the movement of residents between compartments of the susceptible (S), exposed (E), infectious (I), vaccinated (V), immune (R), and deaths (D).

**Figure 2.**
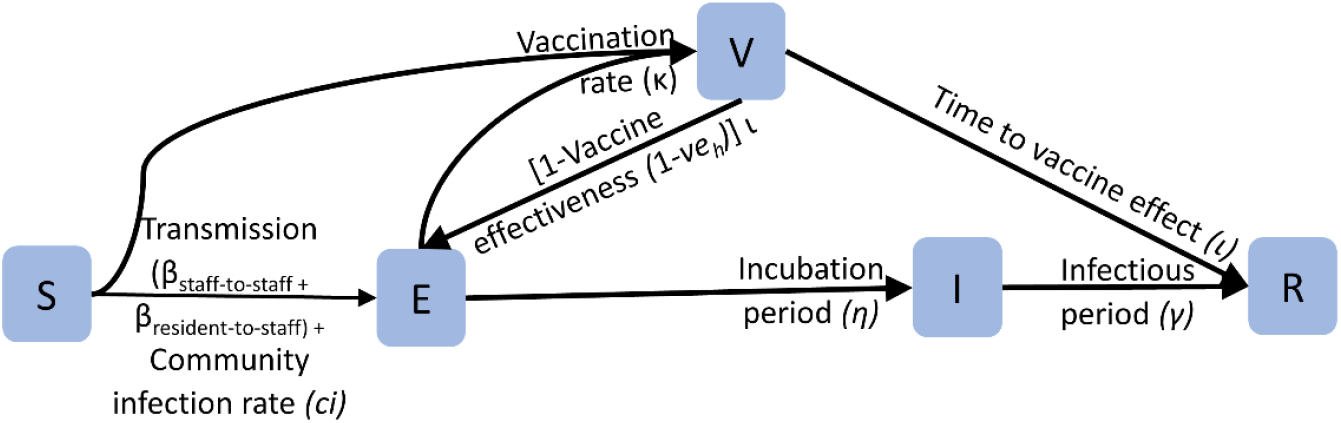
Schematic of the movement of staff between compartments of the susceptible (S), exposed (E), infectious (I), vaccinated (V), and immune (R).

The model parameters governing movement between compartments included both estimated and fixed parameters. The estimated transmission rate parameters, β_resident-to-resident_, β_resident-to-staff_, β_staff-to-staff_, and β_staff-to-resident_ were each multiplied by the number of infected staff (β_staff-to-staff,_ β_staff-to-resident_) or infected residents (β_resident-to-resident_, β_resident-to-staff_) and the number of susceptible staff (β_resident-to-staff,_ β_staff-to-staff_) or susceptible residents (β_resident-to-resident,_ β_staff-to-resident_) to capture the transmission direction. The fixed parameters included η time-to-infectiousness, γ recovery rate, κ vaccination rate, *ve* vaccine effectiveness, ι immunization rate through vaccination, ρ death rate, ρ_C_ COVID-specific death rate, τ discharge rate, *ci* community infection prevalence and σ rate of waning immunity. Parameters governing the distribution of new admissions among the compartments included *ci* community infection prevalence, *imm* proportion of people 65 and older in the county with previous vaccination, and *cvax* vaccination rate among people 65 and older. The values used for each parameter of each epidemic phase and each categorization scheme model can be found in Tables S1-S5 in the Supplement.

### Sampling distributions

To fit the I, V, and D compartments to data, sampling distributions were used with parameters informed by the fitted compartments. Weekly case counts, vaccinations, and deaths were each assigned a Binomial distribution with *N* total residents or staff and a probability from compartment I, V, or D. Weekly case counts were multiplied by a parameter for the proportion of cases detected, p_detect_, which was also estimated by the model. In estimating transmission events per week (β) and p_detect_, parameters were assigned prior distributions based on information available: the βs were assigned an exponential distribution with a rate of 0.5, and p_detect_ was assigned a Beta distribution with a scale of 30 and a shape of one, truncated on the lower end at 70% detection (Supplement).

### Parameter estimation

Parameters were estimated using Bayesian inversion methods related to those previously used in modeling the COVID-19 epidemic and which allow for prior knowledge about parameters to be used and uncertainty around estimates to be produced (6-8). Models were run in the Stan computational platform through R version 4.0.4 and packages detailed in the Supplement (9, 10). Each model was run for at least 60,000 iterations across four chains. To limit autocorrelation in the estimate distributions, the last 30,000 iterations were thinned to every 10^th^ iteration in each chain. Convergence on the final estimates was determined through chain visualization and a Gelman-Rubin convergence statistic of 1.01 or less (11). Details on the post-estimation assessments of model fit and parameter sensitivity can be found in the Supplement. To compare transmission rate estimates between NH categories, an overall posterior distribution of transmission events per week for each category was produced by summing across pathways and averaging the three time periods.

### Credible intervals

Bayesian credible intervals were calculated from each transmission estimate’s posterior probability distribution; unlike the frequentist’s confidence interval, credible intervals provide the expected probability that the estimand is greater than or less than a given value (12). To compare rate estimates between transmission pathways within epidemic phases and NH types, the area of the posterior distributions outside the comparator’s 95% credible interval was calculated as the probability of a significant difference.

### Human Subject Determination and Consent

This activity was reviewed by CDC, deemed not research, and was conducted consistent with applicable federal law and CDC policy.^§^

## RESULTS

### Data summary

Average weekly case incidence was highest during the Omicron phase at 43 per 1,000 residents and 40 per 1,000 staff and lowest during Delta with <6 per 1,000 among both residents and staff (Fig. 3A). The average weekly new vaccinations were highest during Delta, peaking at nine per 1,000 residents and 24 per 1,000 staff and decreased to an average of 0.4 per 1,000 residents and five per 1,000 staff during Omicron (Fig. 3B). The average weekly death rates among residents were highest during pre-Delta, averaging eight per 1,000 resident-weeks (Fig. 3B).

**Figure 3.**
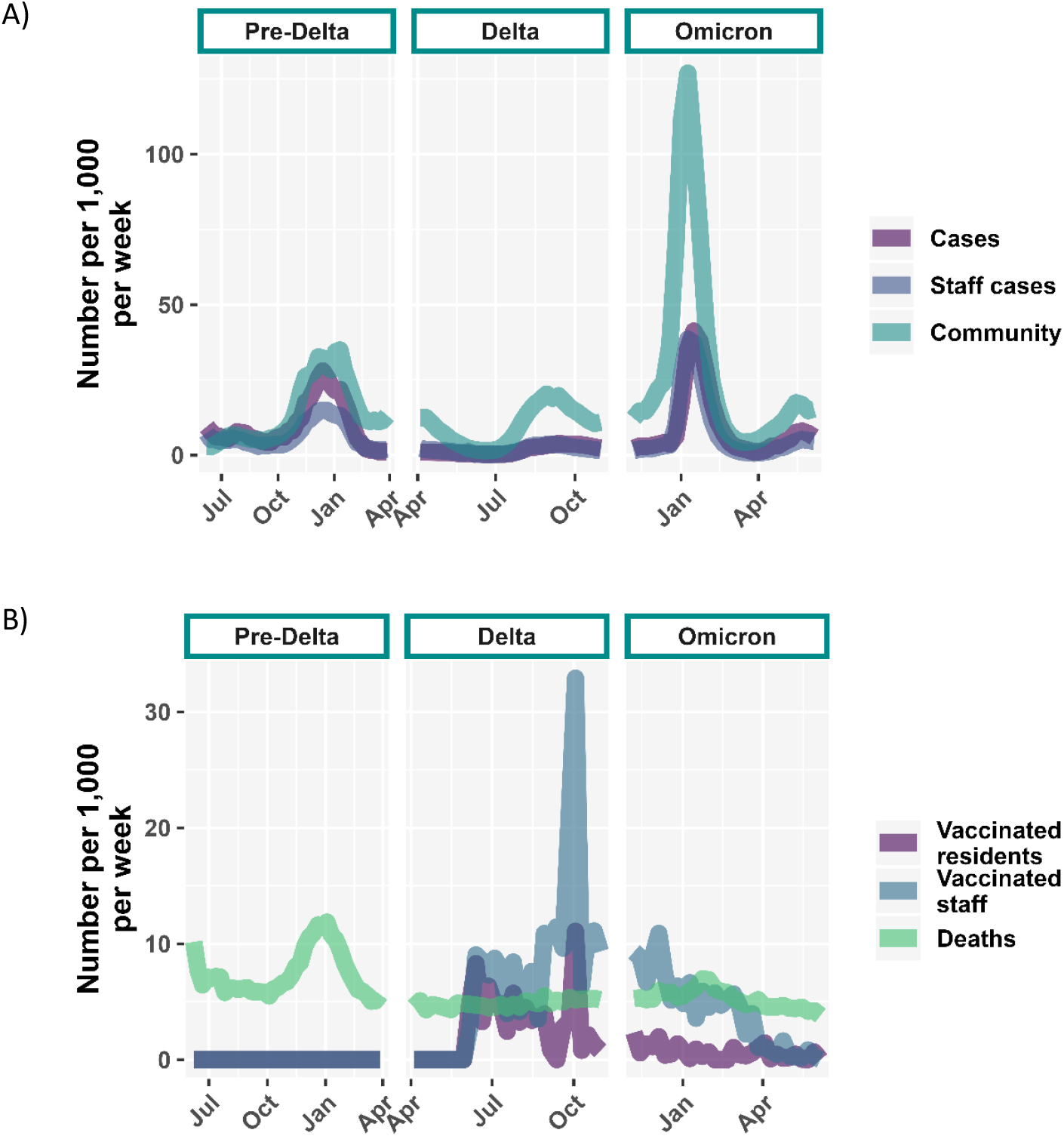
A) Trends in the number of COVID-19 cases among residents (purple), staff (blue), and the community (green) per 1,000 people per week categorized by the dominant variant of the time period. B) Trends in the number of vaccinated residents (purple) and staff (blue), and resident deaths (green) per 1,000 people per week categorized by the dominant variant of the time period.

### Parameter estimation and model fit

Across categorization schemes, average transmission rates were high during the pre-Delta and Omicron periods, and the resident-to-resident or staff-to-resident transmission pathways typically were highest. Across categorizations, model fit was poor during the Delta period due to the low case incidence (Supplement Table S7). In sensitivity analyses, a flat prior distribution for the proportion of cases detected resulted in the largest difference of greater than three standard deviations in the estimated transmission rates (Supplement Fig. S1).

#### By nursing home size

Comparing the median transmission events per week between the largest 1% of NH (>299 beds) and all others (<300 beds), the median rate was 17% higher in NH with <300 beds and the overall probability of a significantly higher transmission rate was 51% (Table 1). When comparing transmission pathways and time periods within each NH category, the highest median transmission rate in NH with <300 beds was during the pre-Delta period from staff to residents (median: 0.67, 95% credible interval (CrI): 0.03-1.65), and in NH with >299 beds during Omicron between residents (median: 0.66, 95% CrI: 0.13-0.93) (Fig. 4).

**Table 1.**
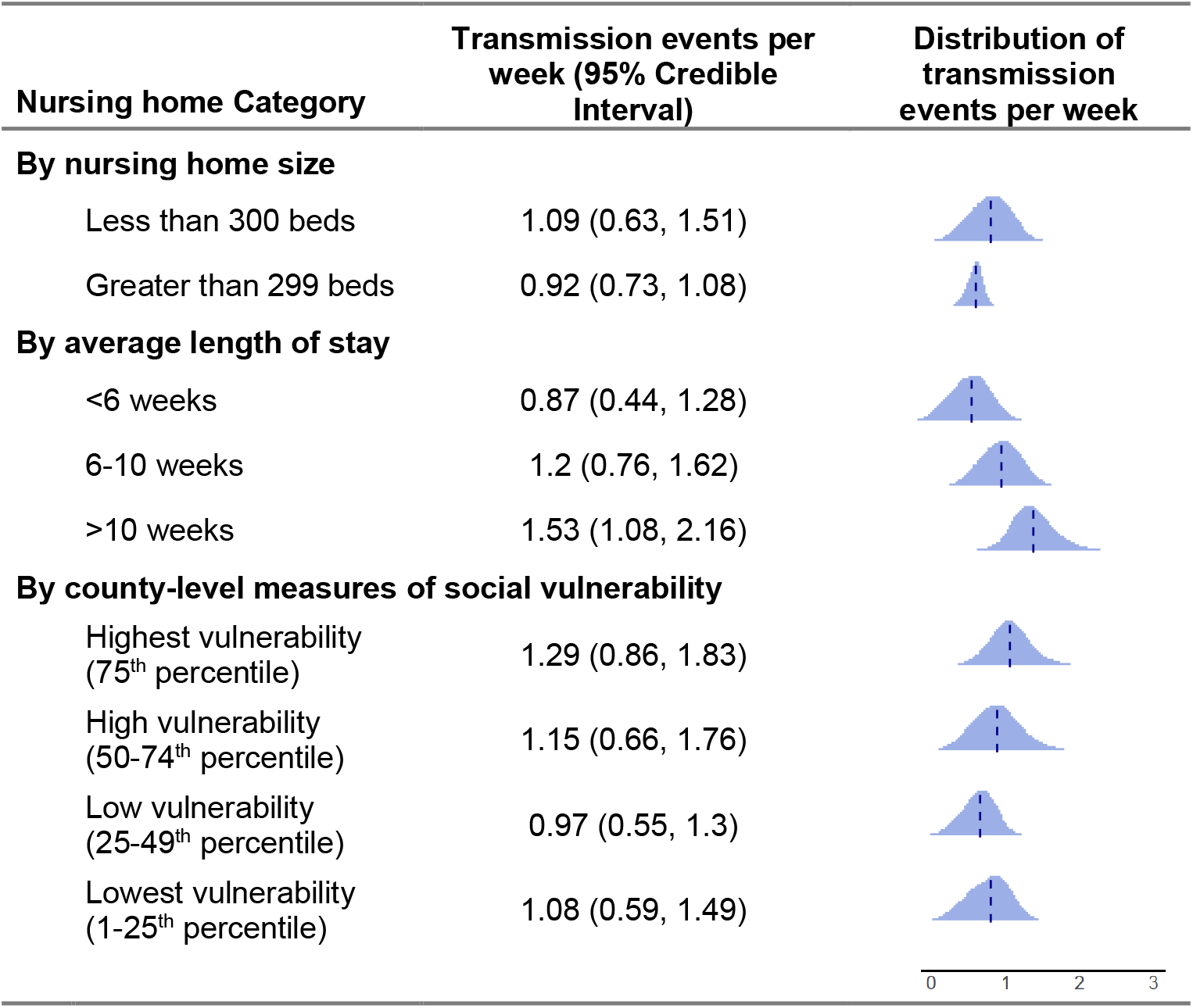
Estimated median and distributions of the transmission events per week for each nursing home category, summed across transmission pathways and averaged over the three time periods.

| Nursing home Category | Transmission events per week (95% Credible Interval) | Distribution of transmission events per week |
| --- | --- | --- |
| **By nursing home size** |  |  |
| Less than 300 beds | 1.09 (0.63, 1.51) |  |
| Greater than 299 beds | 0.92 (0.73, 1.08) |  |
| **By average length of stay** |  |  |
| <6 weeks | 0.87 (0.44, 1.28) |  |
| 6-10 weeks | 1.2 (0.76, 1.62) |  |
| >10 weeks | 1.53 (1.08, 2.16) |  |
| **By county-level measures of social vulnerability** |  |  |
| Highest vulnerability (75<sup>th</sup> percentile) | 1.29 (0.86, 1.83) |  |
| High vulnerability (50-74<sup>th</sup> percentile) | 1.15 (0.66, 1.76) |  |
| Low vulnerability (25-49<sup>th</sup> percentile) | 0.97 (0.55, 1.3) |  |
| Lowest vulnerability (1-25<sup>th</sup> percentile) | 1.08 (0.59, 1.49) |  |

**Figure 4.**
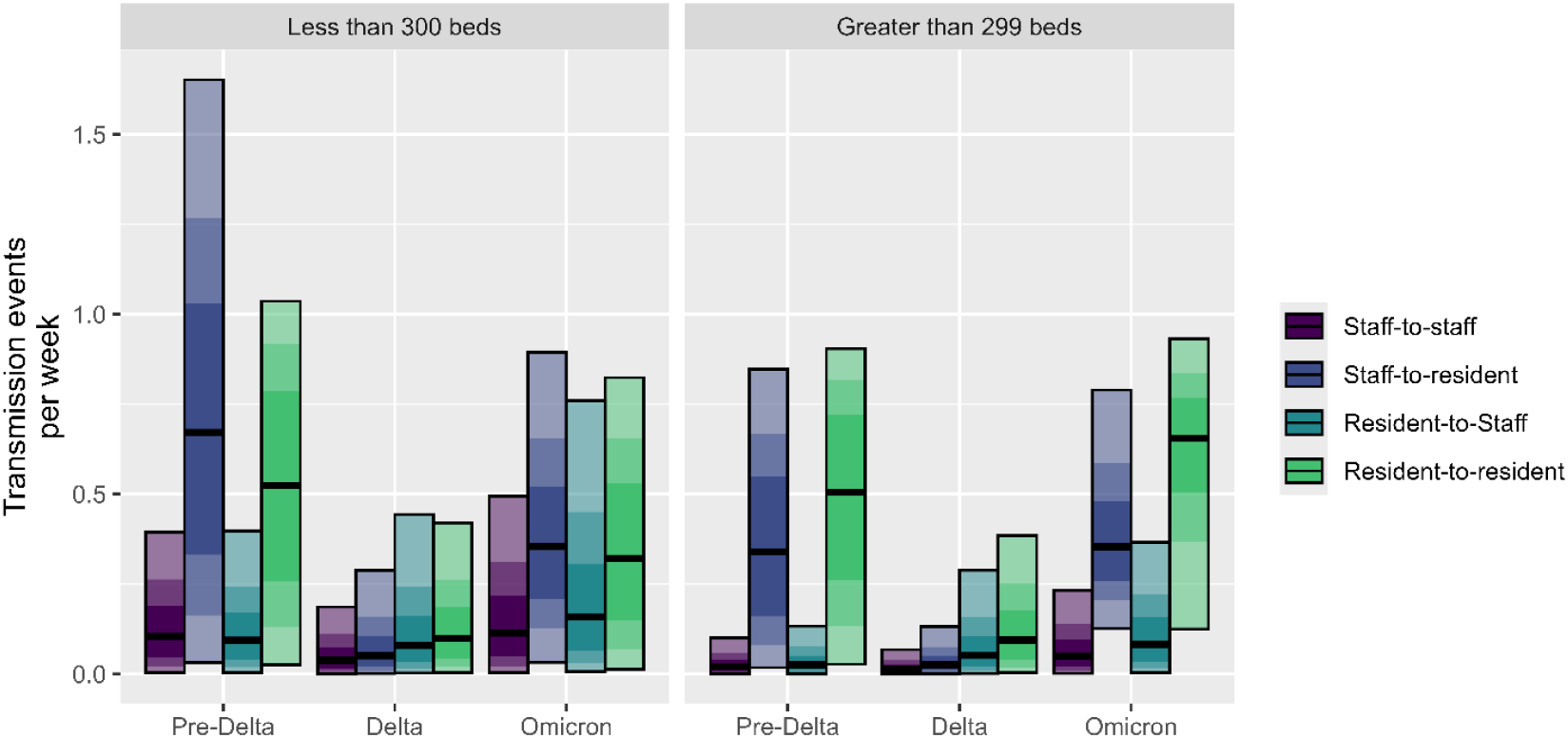
Estimates for the number of transmission events per week between staff (red-purple), from staff to residents (purple), from residents to staff (blue), and between residents (green) for facilities categorized by the number of beds. The black horizontal line indicates the median estimate for each transmission route. The shade of each bar corresponds to the credible interval (CrI) width for the estimate with the darkest shade containing the most credible of the estimates; the lightest shade is the 95% CrI, the next darkest shade is the 75% CrI, and the darkest shade is the 50% CrI.

#### By average length of stay

Comparing the median transmission rates between NH categorized by the median resident LOS, the rate in NH with a median LOS >10 weeks was 74% higher than the rate in NH with a median LOS <6 weeks and 26% higher than the rate in NH with a median LOS 6-10 weeks (Table 1). The probability of a significantly higher transmission rate in NH with a median LOS >10 weeks was 88% compared to NH with a median LOS <6 weeks and 36% compared to NH with a median LOS 6-10 weeks (Table 1). When comparing transmission pathways and time periods within NH categories, the highest median transmission rate in NH with average LOS <6 weeks was during pre-Delta from staff to residents (0.47, 95% CrI: 0.02-1.34) and resident to resident (0.47, 95% CrI: 0.02-0.95), in NH with average LOS 6-10 weeks was during pre-Delta from staff to residents (0.68, 95% CrI: 0.03-1.78), and in NH with average LOS >10 weeks was during pre-Delta from staff to residents (0.88, 95% CrI: 0.06-1.85) (Fig. 5).

**Figure 5.**
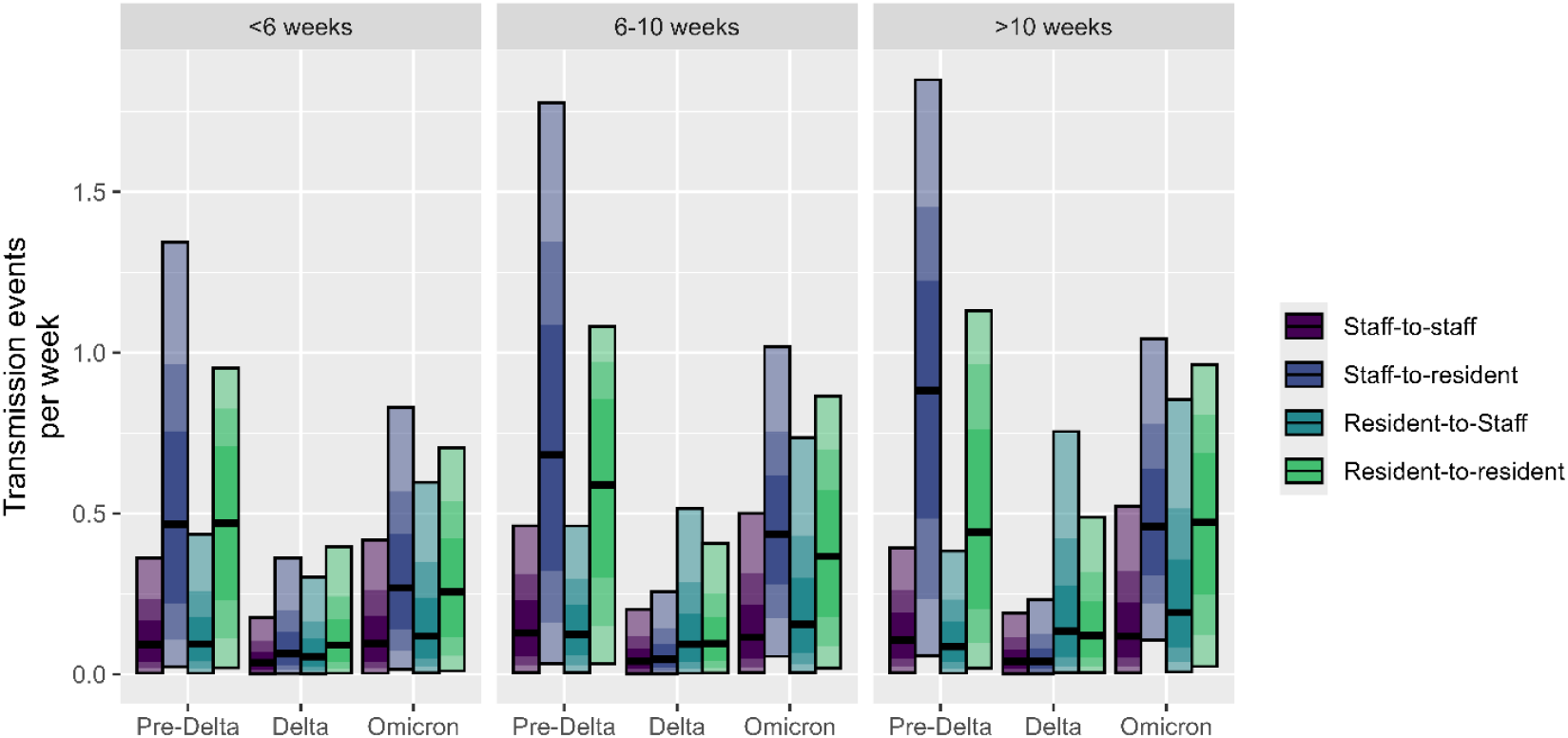
Estimates for the number of transmission events per week between staff (red-purple), from staff to residents (purple), from residents to staff (blue), and between residents (green) for facilities with a median nursing home-wide length of stay of less than six weeks, 6-10 weeks, or greater than 10 weeks. The black horizontal line indicates the median estimate for each transmission route. The shade of each bar corresponds to the credible interval (CrI) width for the estimate with the darkest shade containing the most credible of the estimates; the lightest shade is the 95% CrI, the next darkest shade is the 75% CrI, and the darkest shade is the 50% CrI.

#### By county-level measures of social vulnerability

Comparing the median transmission rates between NH categorized by the SVI of the county in which they were located, the median transmission rate was highest in NH located in the highest vulnerability counties (75^th^ percentile). The largest difference in median transmission rate, 33%, was between NH in the highest SVI quartile (75^th^ percentile) (1.27, 95% CrI: 0.17-1.92) and those in the third (low) SVI quartile (25-49^th^ percentile) (0.95, 95% CrI: 0.15-1.49) (Table 1). The probability of a significantly higher transmission rate in NH in the highest vulnerability counties was 48% compared to those in the third (low) SVI quartile (25-49^th^ percentile) (Table 1). When comparing transmission pathways and time periods within NH categories, transmission was highest in NH located in counties with the highest social vulnerability (75^th^ percentile) during pre-Delta from staff to residents (0.87, 95% CrI: 0.13-1.74), in those with high social vulnerability (50-74^th^ percentile) during pre-Delta from staff to residents (0.77, 95% CrI: 0.09-1.68), in those with low social vulnerability (25-49^th^ percentile) during pre-Delta from resident to resident (0.60, 95% CrI: 0.03-1.00), and in those with lowest social vulnerability (1-25^th^ percentile) during pre-Delta from resident to resident (median: 0.89, 95% CrI: 0.07-1.16) (Fig. 6).

**Figure 6.**
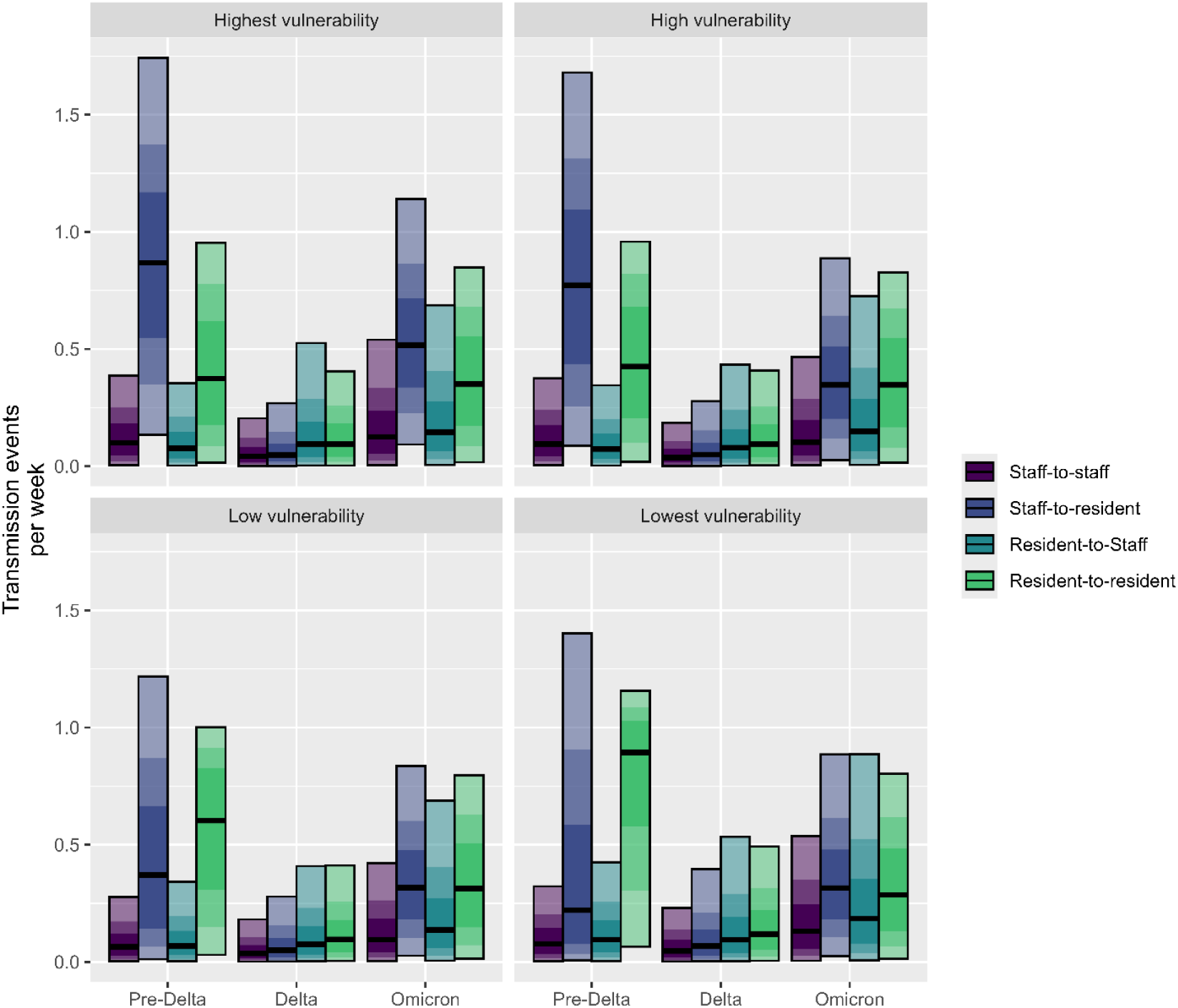
Estimates for the number of transmission events per week between staff (red-purple), from staff to residents (purple), from residents to staff (blue), and between residents (green) for facilities categorized as the highest (76-100th percentile), high (51-75th percentile), low (26-50th percentile), or lowest (0-25th percentile) vulnerability according to the Minority Health Social Vulnerability Index. The black horizontal line indicates the median estimate for each transmission route. The shade of each bar corresponds to the credible interval (CrI) width for the estimate with the darkest shade containing the most credible of the estimates; the lightest shade is the 95% CrI, the next darkest shade is the 75% CrI, and the darkest shade is the 50% CrI.

## DISCUSSION

We estimated the directionality and rate of transmission for SARS-CoV-2 in NH and found that staff-to-resident and resident-to-resident were the dominant pathways of transmission in the earlier part of the pandemic, prior to the Delta variant’s predominance and during the Omicron wave, though not as strongly. When stratifying NH either based on size, the average resident LOS, or the surrounding county’s SVI, we found higher transmission estimates in NH with a longer average resident LOS and located in counties with higher social vulnerability. The dominance of the staff-to-resident and resident-to-resident transmission pathways across categories and time periods indicates where interventions may have the most impact.

The dominant transmission pathways during each epidemic phase likely were influenced by viral prevalence in the surrounding community, NH visitation, infection prevention and control (IPC) practices, and immunity in the resident and staff populations. In March 2020, the Centers for Medicare and Medicaid Services recommended NH restrict outside visitation (13). As a result, resident community contact was reduced, so viral introductions into the resident population would primarily have been though interactions with staff who had regular close contact and prolonged hands-on care with residents while also interfacing with the community. During this time, the pre-Delta phase, we found the staff-to-resident transmission pathway to be dominant for most categories (Fig. 4, 5 & 6). However, this transmission pathway also exhibited a wide distribution of values in the 95% credible interval, which may reflect variation in IPC practices that were not modeled for lack of data. Completion of the two-dose vaccination series among residents and staff began to increase in May 2021, which, in addition to any natural immunity acquired during the pre-Delta phase, likely contributed to lower transmission during the Delta phase. Visitation guidelines were relaxed in November 2021, as well as some masking requirements in NH, potentially allowing more introduction of the virus from the community through visitors before and during Omicron (13). We found the resident-to-resident transmission pathway, in addition to the staff-to-resident pathway, to be dominant during this time.

Previous efforts to estimate SARS-CoV-2 transmission rates have typically focused on community transmission (14, 15) with estimates ranging from 0.7 to 9.3 transmission events per week, but given NH residents’ vulnerability and their regular interactions with staff, estimates for this setting would likely differ from estimates for the community. Through an international meta-analysis, Thompson et al. (16) estimated transmission risk in varying settings, including healthcare (limited to hospitals). The authors found that, on average, 3.6 secondary cases per case could be expected in a healthcare setting, which was higher than in other workplaces (1.9) but lower than within households (21.1), highlighting how much transmission can vary depending on setting (16). Adding another dimension to the complexities of setting-specific transmission, social vulnerabilities are important to explore in disease transmission to identify risk disparities. A previous study that evaluated risk factors for COVID-19 and indicators for transmission risk in NH found larger NH size, urban location, and a higher percentage of African American residents to be associated with case occurrence (2). We found differences in the dominant transmission pathway during pre-Delta between counties with higher and lower social vulnerabilities, with resident-to-resident transmission dominating less vulnerable counties and staff-to-resident transmission dominating higher vulnerability counties. This is in contrast to the Omicron and Delta phases, where the estimated number of transmission events per week was similar across pathways, potentially due to decreased transmission overall given the immunity that had accumulated in residents and the surrounding population, as well as the effects of potential improvements in IPC practices and increased resources and support (17). The highest transmission pathway estimate of all the time periods and social vulnerability categories was between residents in counties in the 0-25^th^ percentile of SVI. However, this does not indicate that transmission was highest in these counties, but rather that this specific transmission pathway accounted for most of the transmission in the NH within these counties. More complete information on resident characteristics would allow for a more detailed analysis of the differences in transmission in individual NH by sociodemographic factors.

In conducting this analysis, we encountered some limitations and were required to make simplifying assumptions. First, the sensitivity analysis revealed that the proportion of cases detected was influential in the estimation of transmission parameters. We allowed the proportion of cases detected to be estimated by the model but constrained detection to be at least 70% of cases. During the study period, testing protocols differed by NH due to constraints on resource and implementation, and there was no systematic information about testing frequency that could have informed the parameter. However, the model fit to the observed data was best when using the constrained proportion detected (Supplement Fig. S1). Second, we were unable to account for the potential introduction of the virus into NH via visitation from members of the community for lack of visitation rates in NH, which changed at the national and NH-level over the study period. Although there is a lack of evidence on compliance of visitor restrictions among NHs, Graham, et.al. and Blackman, et. al. demonstrated that the virus still was introduced into NHs with visitor restrictions (18, 19). Viral introduction by visitors could account for a portion of the estimated transmission from staff to residents given that both are the result of exposures in the community outside of the NH; however, without information about visitation, we cannot say how small or large that portion would be. A third limitation of this study is the inability to account for staff turnover (e.g., absence due to illness, leaving a position, etc.) and shortages without a comprehensive data source of individual staff hours, which would affect the attributable portion of the transmission coming from staff.

We have constructed a model framework to estimate the transmission dynamics of SARS-CoV-2 in nursing homes. The dominance of the staff-to-resident and resident-to-resident transmission pathways across the NH categorization schemes evaluated here indicates that interventions that focus on interrupting these transmission pathways may have the most impact.

## Data Availability

Publicly-available data sources are listed in the methods section. Any supplemental data can be made available upon reasonable request to the authors.

## DISCLAIMER

The findings and conclusions in this report are those of the author(s) and do not necessarily represent the views of the Centers for Disease Control and Prevention.

## Footnotes

§ See e.g., 45 C.F.R. part 46.102(l)(2), 21 C.F.R. part 56; 42 U.S.C. §241(d); 5 U.S.C. §552a; 44 U.S.C. §3501 et seq.

