## Supplement for "Estimating transmission pathways of COVID-19 in U.S. nursing homes"

Table S6. The probability of higher transmission comparing between the dominant transmission pathway (grey) for each nursing home type and time period and all other transmission pathways. . 5

### Model structure

The STAN model code can be found at the end of the document.

#### Resident compartment group

$$\frac{dS}{dt} = admissions * (1-ci-cvax-imm) - IS\,\beta_{resident\text{-}resident} - I_hS\,\beta_{HCP\text{-}resident} - \kappa S - \rho S - \tau S$$

$$\frac{dE}{dt} = IS\,\beta_{resident\text{-}resident} + I_hS\,\beta_{HCP\text{-}resident} + (1\text{-}ve) * \iota V - \eta E - \kappa E - \rho E - \tau E$$

$$\frac{dI}{dt} = \eta E - \gamma I - \rho CI + admissions * ci$$

$$\frac{dV}{dt} = \kappa(S+E) - \iota V - (1\text{-}ve) * \iota V - \rho V - \tau V + admissions * cvax$$

$$\frac{dR}{dt} = \gamma I + \iota V - \rho R - \tau R + admissions * imm$$

$$\frac{dD}{dt} = \rho(S+E+V+R) + \rho_C I - D$$

$$Admissions = \tau(S+E+V+R) + D$$

### Healthcare personnel compartment group

$$\frac{dS_h}{dt} = \sigma R_h - I_h S_h \beta_{HCP\text{-}to\text{-}HCP} - IS_h \beta_{resident\text{-}to\text{-}HCP} - ciS_h - \kappa_h S_h$$

$$\frac{dE_h}{dt} = I_h S_h \beta_{HCP\text{-}to\text{-}HCP} + IS_h \beta_{resident\text{-}to\text{-}HCP} + (1\text{-}ve_h) * \iota V_h + ciS_h - \eta E_h - \kappa_h E_h$$

$$\frac{dI_h}{dt} = \eta E_h - \gamma I_h$$

$$\frac{dV_h}{dt} = \kappa_h (S_h + E_h) - \iota V_h - (1\text{-}ve_h) * \iota V_h$$

$$\frac{dR_h}{dt} = \gamma I_h + \iota V_h - \sigma R_h$$

### Sampling distributions

Resident case counts ~ Binomial(N, $I$ * Proportion_detected)

HCP case counts ~ Binomial ($N_h$, $I_h$ * Proportion_detected)

Vaccinated resident counts ~ Binomial (N, $V$)

Vaccinated HCP counts ~ Binomial ($N_h$, $V_h$)

Resident death counts ~ Binomial (N, $D$)

### Prior distributions

$\beta_{resident\text{-}resident}$ ~ Exponential(.5)

$\beta_{resident\text{-}to\text{-}HCP}$ ~ Exponential (.5)

$\beta_{HCP\text{-}to\text{-}HCP}$ ~ Exponential (.5)

$\beta_{HCP\text{-}resident}$ ~ Exponential (.5)

Proportion_detected ~ Beta(10, 1)

### Model Parameters

#### Table S1. Parameters for the model shared across categorization schemes

| Parameter | Description | Pre-Delta | Delta | Omicron | Reference |
|---|---|---|---|---|---|
| ve | Vaccine effectiveness against infection - residents | - | 0.6 | 0.5 | [1, 2] |
| $ve_h$ | Vaccine effectiveness against infection - staff | - | 0.8 | 0.5 | [3, 4] |
| ci | Community infection rate per 1,000 | 9.3 | 9.5 | 14.6 | [5] |
| cvax | Community 65+ vaccination rate per 1,000 | - | 3.4 | 1.1 | [6] |
| imm | Community 65+ proportion immune | 0.0 | 0.4 | 0.1 | [5] |
| $\eta$ | Incubation period (weeks) | 0.7 | 0.7 | 0.7 | [7] |
| $\gamma$ | Infectious period (weeks) | 1.0 | 1.0 | 1.0 | [8] |
| $\sigma$ | Period of immunity (weeks) | 25.0 | 25.0 | 25.0 | [9, 10] |
| $\iota$ | Time to vaccine effect for 2-dose series (weeks) | - | 4.0 | 4.0 | [11] |

#### Table S2. Parameters for the model with facilities categorized by size based on the number of reported beds

| Parameter | Description | Less than 300 beds | | | Largest 1% (Greater than 299 beds) | | | Reference |
|---|---|---|---|---|---|---|---|---|
| | | Pre-Delta | Delta | Omicron | Pre-Delta | Delta | Omicron | |
| $\kappa$ | Vaccination rate per 1,000 - residents | - | 2.6 | 0.6 | - | 2.7 | 0.7 | [12] |
| $\kappa_h$ | Vaccination rate per 1,000 - staff | - | 5.8 | 4.5 | - | 7.9 | 3.8 | [12] |
| $\rho$ | Death rate per 1,000 | 8.3 | 5.1 | 5.4 | 6.6 | 4.6 | 5.1 | [12] |
| $\rho_C$ | COVID death rate per 1,000 | 2.5 | 0.3 | 0.4 | 1.6 | 0.2 | 0.3 | [12] |
| $\tau$ | Length of stay (weeks) | 6.2 | 5.6 | 5.8 | 14.3 | 13.3 | 13.4 | Estimated from Unpublished CMS Minimum dataset |

### Table S3. Parameters for the model with facilities categorized by average resident length of stay

| Parameter | Description | <6 weeks | | | 6-10 weeks | | | >10 weeks | | | Reference |
|---|---|---|---|---|---|---|---|---|---|---|---|
| | | Pre-Delta | Delta | Omicron | Pre-Delta | Delta | Omicron | Pre-Delta | Delta | Omicron | |
| $\kappa$ | Vaccination rate per 1,000 - residents | - | 2.9 | 0.8 | - | 2.6 | 0.5 | - | 2.1 | 0.5 | [12] |
| $\kappa_h$ | Vaccination rate per 1,000 - staff | - | 5.9 | 3.7 | - | 5.8 | 4.8 | - | 5.9 | 4.9 | [12] |
| $\rho$ | Death rate per 1,000 | 9.6 | 6.2 | 6.6 | 8.5 | 5.0 | 5.2 | 6.5 | 3.8 | 4.2 | [12] |
| $\rho_C$ | COVID death rate per 1,000 | 2.6 | 0.3 | 0.5 | 2.6 | 0.3 | 0.4 | 2.1 | 0.2 | 0.3 | [12] |
| $\tau$ | Length of stay (weeks) | 3.6 | 3.5 | 3.6 | 7.5 | 6.4 | 6.8 | 17.3 | 13.0 | 14.1 | Estimated from Unpublished CMS Minimum dataset |

### Table S4. Parameters for the model with facilities categorized by the surrounding county's social vulnerability index

| Parameter | Description | Highest vulnerability | | | High vulnerability | | | Low vulnerability | | | Lowest vulnerability | | | Reference |
|---|---|---|---|---|---|---|---|---|---|---|---|---|---|---|
| | | Pre-Delta | Delta | Omicron | Pre-Delta | Delta | Omicron | Pre-Delta | Delta | Omicron | Pre-Delta | Delta | Omicron | |
| $\kappa$ | Vaccination rate per 1,000 - residents | - | 3.1 | 0.8 | - | 2.6 | 0.6 | - | 2.3 | 0.5 | - | 1.7 | 0.3 | [12] |
| $\kappa_h$ | Vaccination rate per 1,000 - staff | - | 6.3 | 4.8 | - | 6.1 | 4.3 | - | 5.6 | 4.4 | - | 4.7 | 3.9 | [12] |

| Parameter | Description | Highest vulnerability | | | High vulnerability | | | Low vulnerability | | | Lowest vulnerability | | | Reference |
|---|---|---|---|---|---|---|---|---|---|---|---|---|---|---|
| | | Pre-Delta | Delta | Omicron | Pre-Delta | Delta | Omicron | Pre-Delta | Delta | Omicron | Pre-Delta | Delta | Omicron | |
| $\rho$ | Death rate per 1,000 | 7.3 | 4.1 | 4.2 | 8.1 | 5.1 | 5.3 | 9.0 | 5.8 | 6.2 | 10.3 | 6.5 | 7.2 | [12] |
| $\rho_C$ | COVID death rate per 1,000 | 2.5 | 0.2 | 0.3 | 2.4 | 0.3 | 0.4 | 2.5 | 0.3 | 0.5 | 2.7 | 0.3 | 0.5 | [12] |
| $\tau$ | Length of stay (weeks) | 7.1 | 6.0 | 6.4 | 6.4 | 5.5 | 5.7 | 6.0 | 5.3 | 5.4 | 6.2 | 5.4 | 5.6 | Estimated from Unpublished CMS Minimum dataset |

### Transmission estimate comparisons

Table S6. The probability of higher transmission comparing the credible intervals between the dominant transmission pathway (grey) for each nursing home type and time period and all other transmission pathways. The darker shade of blue indicates increasing probability of significantly higher transmission in dominant transmission pathway.

| | | | Comparator pathway | | | |
|---|---|---|---|---|---|---|
| **Nursing home Category** | **Outbreak period** | **Dominant pathway** | **Staff-to-staff** | **Resident-to-Staff** | **Resident-to-resident** | **Staff-to-resident** |
| **By nursing home size** | | | | | | |
| Less than 300 beds | Pre-Delta | Staff-to-resident | 70 | 70 | 25 | |
| | Delta | Resident-to-resident | 25 | 2 | | 10 |
| | Omicron | Staff-to-resident | 28 | 6 | 4 | |
| Greater than 299 beds | Pre-Delta | Resident-to-resident | 91 | 88 | | 9 |
| | Delta | Resident-to-resident | 62 | 8 | | 37 |
| | Omicron | Resident-to-resident | 94 | 88 | | 21 |
| **By average length of stay** | | | | | | |
| <6 weeks | Pre-Delta | Resident-to-resident | 61 | 53 | | 0 |

| | | | Comparator pathway | | | |
|---|---|---|---|---|---|---|
| **Nursing home Category** | **Outbreak period** | **Dominant pathway** | **Staff-to-staff** | **Resident-to-Staff** | **Resident-to-resident** | **Staff-to-resident** |
| | Delta | Resident-to-resident | 24 | 7 | | 4 |
| | Omicron | Staff-to-resident | 27 | 11 | 6 | |
| | Pre-Delta | Staff-to-resident | 65 | 65 | 25 | |
| 6-10 weeks | Delta | Resident-to-resident | 20 | 1 | | 12 |
| | Omicron | Staff-to-resident | 40 | 14 | 7 | |
| | Pre-Delta | Staff-to-resident | 80 | 80 | 32 | |
| >10 weeks | Delta | Resident-to-Staff | 38 | | 9 | 31 |
| | Omicron | Resident-to-resident | 45 | 9 | | 1 |
| **By county-level measures of social vulnerability** | | | | | | |
| Highest vulnerability | Pre-Delta | Staff-to-resident | 85 | 87 | 43 | |
| | Omicron | Staff-to-resident | 46 | 28 | 14 | |
| High vulnerability | Pre-Delta | Staff-to-resident | 79 | 81 | 35 | |
| | Delta | Resident-to-resident | 24 | 2 | | 10 |
| Low vulnerability | Pre-Delta | Resident-to-resident | 77 | 72 | | 0 |
| | Delta | Resident-to-resident | 25 | 3 | | 10 |
| | Omicron | Staff-to-resident | 33 | 7 | 3 | |
| Lowest vulnerability | Pre-Delta | Resident-to-resident | 87 | 82 | | 0 |
| | Delta | Resident-to-resident | 24 | 2 | | 6 |
| | Omicron | Staff-to-resident | 19 | 2 | 4 | |

### Post estimation assessment methods

*Posterior predictive checking.* Posterior prediction simulations were produced from the model in order to evaluate the model fit by removing the sampling distributions and setting the transmission and $p_{detect}$

parameters to their estimated values. The average weekly case counts were used as a test quantity to assess fit numerically, with the ideal scenario of the value falling around the 50th percentile of the posterior prediction. The difference between the 50th percentile from the average weekly case counts was calculated as the Posterior Prediction Deviation (PPD) with a lower value indicating a better fit to the data. A deviation of 25% or less from the 50th percentile was considered a reasonable fit, corresponding to, at most, a deviation of 9 cases per person-week.

### Table S7. Posterior predictive check for each category and time period model

| Facility Category | Time period | Posterior Prediction Deviation (%) |
|---|---|---|
| **By facility size** | | |
| Less than 300 beds | Pre-Delta | 7.8 |
| | Delta | 37.3 |
| | Omicron | 17.9 |
| Greater than 299 beds | Pre-Delta | 18.0 |
| | Delta | 28.1 |
| | Omicron | 27.4 |
| **By average length of stay** | | |
| <6 weeks | Pre-Delta | 7.5 |
| | Delta | 27.2 |
| | Omicron | 8.5 |
| 6-10 weeks | Pre-Delta | 13.8 |
| | Delta | 31.4 |
| | Omicron | 0.8 |
| >10 weeks | Pre-Delta | 12.5 |
| | Delta | 37.1 |
| | Omicron | 1.7 |
| **By county-level measures of social vulnerability** | | |
| High vulnerability | Pre-Delta | 9.4 |
| | Delta | 30.4 |
| | Omicron | 2.2 |

| Facility Category | Time period | Posterior Prediction Deviation (%) |
|---|---|---|
| Low vulnerability | Pre-Delta | 3.2 |
| | Delta | 30.0 |
| | Omicron | 5.5 |
| Highest vulnerability | Pre-Delta | 13.7 |
| | Delta | 31.1 |
| | Omicron | 2.3 |
| Lowest vulnerability | Pre-Delta | 5.1 |
| | Delta | 35.4 |
| | Omicron | 17.7 |

*Sensitivity analysis.* To evaluate the influence of the prior distributions and fixed parameter values on the estimates of the transmission parameters (number of transmission events per week), the models were adjusted to produce estimates under alternative scenarios: flat prior distributions for the transmission and detection parameters, a flat prior distribution for the detection parameter only, and "extreme" values for the time-to-effect for vaccination, the length of immunity, and the vaccine effectiveness. The posterior estimates of the transmission parameters were compared to the original estimates by calculating the number of standard deviations from the original estimates.

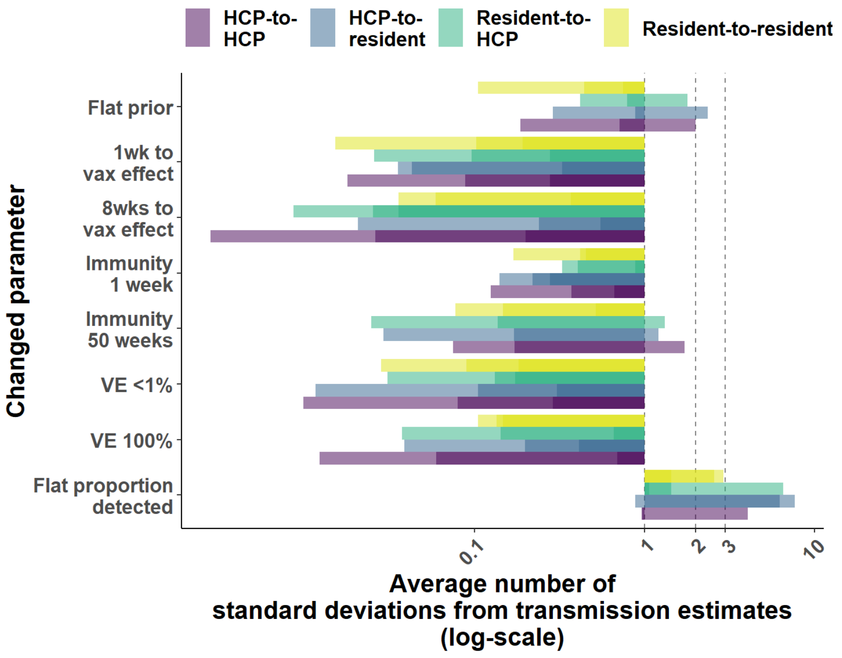

Figure S1. Sensitivity analysis of the fixed parameters and prior distributions assigned in the model. Sensitivity was measured as the average number of standard deviations the transmission estimates were from the estimates produced by the model presented in the paper for facility size. The colors correspond to the estimated transmission pathway and the darker shades represent agreement among the models for each time period. Note: HCP is healthcare provider, i.e., staff.

### R packages used in the analysis

- Rstan 2.21.5
- tidyverse 1.3.0
- tidybayes 3.0.2

### STAN Model code

```
functions {
  real[] sir(real t, real[] y, real[] theta,
          real[] x_r, int[] x_i) {
    //Compartment mapping
    real S = y[1];
    real E = y[2];
    real I = y[3];
    real V = y[4];
    real R = y[5];
    real D = y[6];
    real Sh = y[7];
    real Eh = y[8];
    real Ih = y[9];
    real Vh = y[10];
    real Rh = y[11];
    //Fixed value mapping
    real rhoC = x_r[1]; // death rate among residents - E and I and Ev
    real rho = x_r[2]; // death rate among residents - S, V, and R
    real kappa = x_r[3]; // vax rate in residents
```

```
real kappah = x_r[4]; // vax rate in HCW
real sigma = x_r[5]; // 1/period of immunity
real tau = x_r[6]; // 1/length of stay
real eta = x_r[7]; // 1/incubation period
real comminf = x_r[8];
real ve = x_r[9]; //vaccine effectiveness
real veh = x_r[10]; //vaccine effectiveness
real popvax = x_r[11]; //weekly vax'd proportion in 65+ general population
real prtx65pop = x_r[12]; //prop 65+ vaxd in the previous 4-20 weeks
real gamma = x_r[13]; // 1/infectious period
real iota = x_r[14]; // 1/vax effect period
//Estimated parameter mapping
real betar2r = theta[1]; // transmission resident to resident
real betar2h = theta[2]; // transmission resident to HCW
real betah2h = theta[3]; // transmission HCW to HCW
real betah2r = theta[4]; // transmission HCW to resident

//Compartment equations
///Resident model
real dS_dt = (tau*(S+E+V+R) + D)*(1-comminf-popvax-prtx65pop) - betar2r*I*S - betah2r*Ih*S -
kappa*S - rho*S - tau*S; // term (tau*(S+E+V+R) + D) are all new admissions
real dE_dt = betar2r*I*S + betah2r*Ih*S + (1-ve)*iota*V - eta*E - kappa*E - rho*E - tau*E;
real dI_dt = eta*E - gamma*I - rhoC*I + (tau*(S+E+V+R) + D)*comminf;
real dV_dt = kappa*(S+E) - iota*V - (1-ve)*iota*V - rho*V - tau*V + (tau*(S+E+V+R) + D)*popvax;
real dR_dt = gamma*I + iota*V - rho*R - tau*R + (tau*(S+E+V+R) + D)*prtx65pop;
real dD_dt = rho*(S+E+V+R) + rhoC*I - D;
///HCW model
real dSh_dt = sigma*Rh - betah2h*Ih*Sh - betar2h*I*Sh - comminf*Sh - kappah*Sh;
real dEh_dt = betah2h*Ih*Sh + betar2h*I*Sh + (1-veh)*iota*Vh + comminf*Sh - eta*Eh - kappah*Eh;
```

```
    real dIh_dt = eta*Eh - gamma*Ih;

    real dVh_dt = kappah*(Sh+Eh) - iota*Vh - (1-veh)*iota*Vh;

    real dRh_dt = gamma*Ih + iota*Vh - sigma*Rh;

    return {dS_dt, dE_dt, dI_dt, dV_dt, dR_dt, dD_dt, dSh_dt, dEh_dt, dIh_dt, dVh_dt, dRh_dt};

  }

}

data {

  int<lower=1> n_days;

  int N[n_days];

  int Nh[n_days];

  real t0;

  real y0[11];

  real ts[n_days];

  int cases[n_days];

  int infhcw[n_days];

  int vax[n_days];

  int vaxh[n_days];

  int deaths[n_days];

  real rhoC;

  real rho;

  real kappa;

  real kappah;

  real tau;

  real eta;

  real gamma;

  real iota;

  real sigma;
```

```
  real ve;
  real veh;
  real comminf;
  real popvax;
  real prtx65pop;
}
transformed data {
  real x_r[14] = {rhoC, rho, kappa, kappah, sigma, tau, eta, comminf, ve, veh, popvax, prtx65pop, gamma,
iota};
  int x_i[0];
}
parameters {
  real<lower=0, upper=10> betah2r;
  real<lower=0, upper=10> betar2r;
  real<lower=0, upper=10> betar2h;
  real<lower=0, upper=10> betah2h;
  real<lower=.70, upper=1> p_detected;

}
transformed parameters{
  real y[n_days, 11];
  // initial compartement values
  real theta[4] = {betar2r, betar2h, betah2h, betah2r};
  y = integrate_ode_rk45(sir, y0, t0, ts, theta, x_r, x_i);
}
model {
  //priors
  betar2r ~ exponential(.5);
  betar2h ~ exponential(.5);
```

```
  betah2h ~ exponential(.5);
  betah2r ~ exponential(.5);
  p_detected ~ beta(10, 1);

  //sampling distribution
  //col(matrix x, int n) - The n-th column of matrix x. Here the number of infected people
  cases ~ binomial(N, col(to_matrix(y),3)*p_detected);
  infhcw ~ binomial(Nh, col(to_matrix(y),9)*p_detected);
  vax ~ binomial(N, col(to_matrix(y),4));
  vaxh ~ binomial(Nh, col(to_matrix(y),10));
  deaths ~ binomial(N, col(to_matrix(y),6));
}

generated quantities {
  real pred_cases[n_days];
  real pred_vaxd[n_days];
  real pred_death[n_days];
  real pred_infhcw[n_days];
  real pred_vaxh[n_days];
  pred_cases = binomial_rng(N, col(to_matrix(y),3)*p_detected);
  pred_vaxd = binomial_rng(N, col(to_matrix(y),4));
  pred_death = binomial_rng(N, col(to_matrix(y),6));
  pred_infhcw = binomial_rng(N, col(to_matrix(y),9)*p_detected);
  pred_vaxh = binomial_rng(Nh, col(to_matrix(y),10));
}
```

1. Nanduri S, Pilishvili T, Derado G, et al. Effectiveness of Pfizer-BioNTech and Moderna Vaccines in Preventing SARS-CoV-2 Infection Among Nursing Home Residents Before and During

Widespread Circulation of the SARS-CoV-2 B.1.617.2 (Delta) Variant - National Healthcare Safety Network, March 1-August 1, 2021. MMWR Morb Mortal Wkly Rep **2021**; 70(34): 1163-6.
